# Exploring the thermally-controlled fentanyl transdermal therapy to provide constant drug delivery by physics-based digital twins

**DOI:** 10.1101/2023.11.20.23298752

**Authors:** Flora Bahrami, Agnes Psikuta, René Michel Rossi, Alex Dommann, Thijs Defraeye

## Abstract

Transdermal drug delivery is suitable for low-molecular-weight drugs with specific lipophilicity, like fentanyl, which is widely used for cancer-induced pain management. However, fentanyl’s transdermal therapy displays high intra-individual variability. Factors like skin characteristics at application sites and ambient temperature contribute to this variation. In this study, we developed a physics-based digital twin of the human body to cope with this variability and propose better adapted setups. This twin includes an *in-silico* skin model for drug penetration, a pharmacokinetic model, and a pharmacodynamic model. Based on the results of our simulations, applying the patch on the flank (side abdominal area) showed a 15.3% higher maximum fentanyl concentration in the plasma than on the chest. Additionally, the time to reach this maximum concentration when delivered through the flank was 19.8 h, which was 10.3 h earlier than via the upper arm. Finally, this variation led to an 18% lower minimum pain intensity for delivery via the flank than the chest. Moreover, the impact of seasonal changes on ambient temperature and skin temperature by considering the activity level was investigated. Based on our result, the fentanyl uptake flux by capillaries increased by up to 11.8% from an inactive state in winter to an active state in summer. We also evaluated the effect of controlling fentanyl delivery by adjusting the temperature of the patch to alleviate the pain to reach a mild pain intensity (rated three on the VAS scale). By implementing this strategy, the average pain intensity decreased by 1.1 points, and the standard deviation for fentanyl concentration in plasma and average pain intensity reduced by 37.5% and 33.3%, respectively. Therefore, our digital twin demonstrated the efficacy of controlled drug release through temperature regulation, ensuring the therapy toward the intended target outcome and reducing therapy outcome variability. This holds promise as a potentially useful tool for physicians.

**Graphical Abstract:** (Created with BioRender.com and www.flaticon.com)

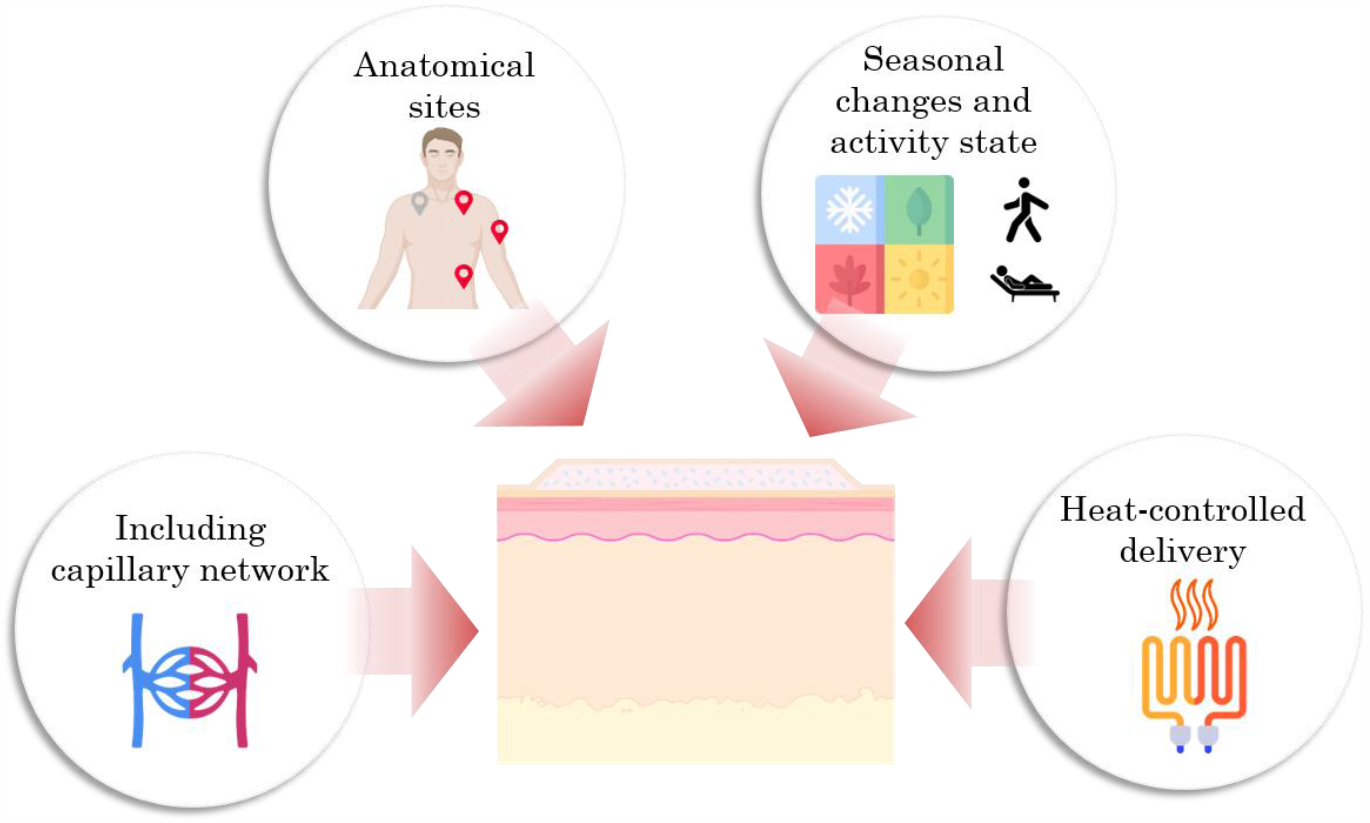

## 1 Introduction

The transdermal delivery system administers drugs through the skin to the body, the largest and outermost organ in the human body (Alkilani, McCrudden, and Donnelly 2015). Transdermal delivery offers benefits, including less fluctuation in drug intake, bypassing first-pass metabolism, and enhancing patient compliance (SHINGADE 2012). On the other hand, it has limitations, such as it is only suitable for low molecular weight potent drugs (Tanwar and Sachdeva 2016). Fentanyl is one of the common drugs administrated transdermally. It is a synthetic opioid that is 50 to 100 times more potent than morphine (Han et al. 2019) and commonly used as an analgesic for cancer-induced pain (Wang et al. 2018). However, fentanyl transdermal therapy shows intra and inter-individual variability (Larsen et al. 2003). This implies that fentanyl trans-dermal therapy leads to different outcomes among patients and even for each patient during the treatment. Various factors contribute to the variability in outcomes following the application of a fentanyl patch. These include differences in skin characteristics at the different application sites on the human body (Thijs Defraeye et al. 2020; Freise et al. 2012; Roy and Flynn 1990), skin temperature (Prodduturi et al. 2010; Zhang et al. 2020) and age, weight, and gender (Dąbrowska et al. 2018). However, the task of fine-tuning the fentanyl dosage for each patient by trial and error is currently the only way to go, despite the mentioned drawbacks.

As a future improvement, computational and mathematical methods could be employed to personalize and adapt transdermal therapy for patients while reducing clinical trial and error methods, which are costly and might put the patient’s health and well-being at risk. Some of these computational methods aim to monitor fentanyl penetration through the skin via molecular dynamics models (Faulkner and de Leeuw 2021; Otto and De Villiers 2013; Rim, Pinsky, and Van Osdol 2009), brick-and-mortar models (Naegel, Heisig, and Wittum 2013), or diffusion models (Anissimov et al. 2013; Bahrami et al. 2023; Bahrami, Rossi, and Defraeye 2022; Thijs Defraeye et al. 2020; Thijs Defraeye, Bahrami, and Rossi 2021; Iordanskii et al. 2000; Walicka and Iwanowska-Chomiak 2018). Additionally, other computational methods focus on the pharmacokinetics and pharmacodynamics model, which aim to predict the drug concentration in plasma by considering the metabolism and eliminations and, eventually, the drug’s effect corresponding to the drug concentration (Bahrami et al. 2023; Bahrami, Rossi, and Defraeye 2022; Björkman 2003; Madden et al. 2019; Pan and Duffull 2019). These models can contribute to evaluating the impact and efficacy of new delivery technologies, such as microneedles (Rajoli et al. 2019; Yan et al. 2022), iontophoresis (Corey et al. 2012; Filipovic et al. 2017), or the application of heat to steer transdermal drug delivery (La Count et al. 2020). Furthermore, some studies explored *in-silico* the impact of anatomical sites on fentanyl delivery (Thijs Defraeye et al. 2020). Also, physics-based digital twins of the human body were developed by combining the transdermal uptake through the *in-silico* model of skin with pharmacokinetics and pharmacodynamics models in order to predict and modify the fentanyl transdermal therapy for virtual patients based on their physiological features and response to the therapy for entire populations of patients (Bahrami et al. 2023; Bahrami, Rossi, and Defraeye 2022). However, these virtual twins of patients for fentanyl therapy have not been used to investigate *in-silico* the impact of novel technologies, such as microneedles, or the impact of thermally-controlled delivery.

In this study, we applied the digital human twin for thermally controlling the fentanyl flux to the body to keep the outcome of the treatment in the acceptable range. For this purpose, we developed a digital twin that includes drug uptake through the *in-silico* skin model, pharmacokinetic, and pharmacodynamic models. For monitoring the drug penetration from the patch through the skin, we developed a new model including different layers of the skin, i.e., stratum corneum, viable epidermis, dermal papillae, dermis, hypodermis, and capillary vessels, to simulate blood flow. Additionally, we took into account the changes in the properties of the skin as the virtual patient implemented the patch on different body sites, including the chest, back, flank, and upper arm. Furthermore, the impact of seasonal changes on the ambient temperature by considering the activity state of the patient on the fentanyl uptake by the skin was studied. In the end, we implemented this temperature dependency of fentanyl penetration in order to steer fentanyl flux to the body in order to control fentanyl concentration in the plasma and, eventually, the pain intensity of the patient.

## 2 Materials and methods

The general approach of this study is to develop a virtual human twin for transdermal therapy, which includes three main blocks of drug uptake, pharmacokinetics, and pharmacodynamics model. To do so, we developed a detailed *in-silico* model of skin, which, besides considering the stratum corneum, viable epidermis, dermis, and hypodermis, includes dermal papillae and capillary networks. Later, this model was used to predict the uptake of fentanyl from the patch by using Fick’s second law and blood flow in the capillaries. Additionally, the Pharmacokinetics model calculates the fentanyl concentration in plasma based on the flux of fentanyl uptake to blood circulation, drug distribution, metabolism, and elimination. Based on the calculated fentanyl concentration in the plasma, the pharmacodynamics model calculates the effect of fentanyl therapy, which is pain relief. Furthermore, within the developed digital twin, we explored the changes in the skin layers in different body sites, such as the chest, back, and upper arm, to evaluate the variation in the therapy outcome between each patch administration. Moreover, we took into account how the ambient temperature of the room the patient resides in affects the fentanyl uptake by considering summer and winter conditions. Later on, we took a step further to implement the impact of heat on fentanyl uptake in order to provide heat-enhanced fentanyl delivery to control the fentanyl concentration and, subsequently, pain intensity in a target range. The overall structure of this study is provided in Figure 1.

**Figure 1.**
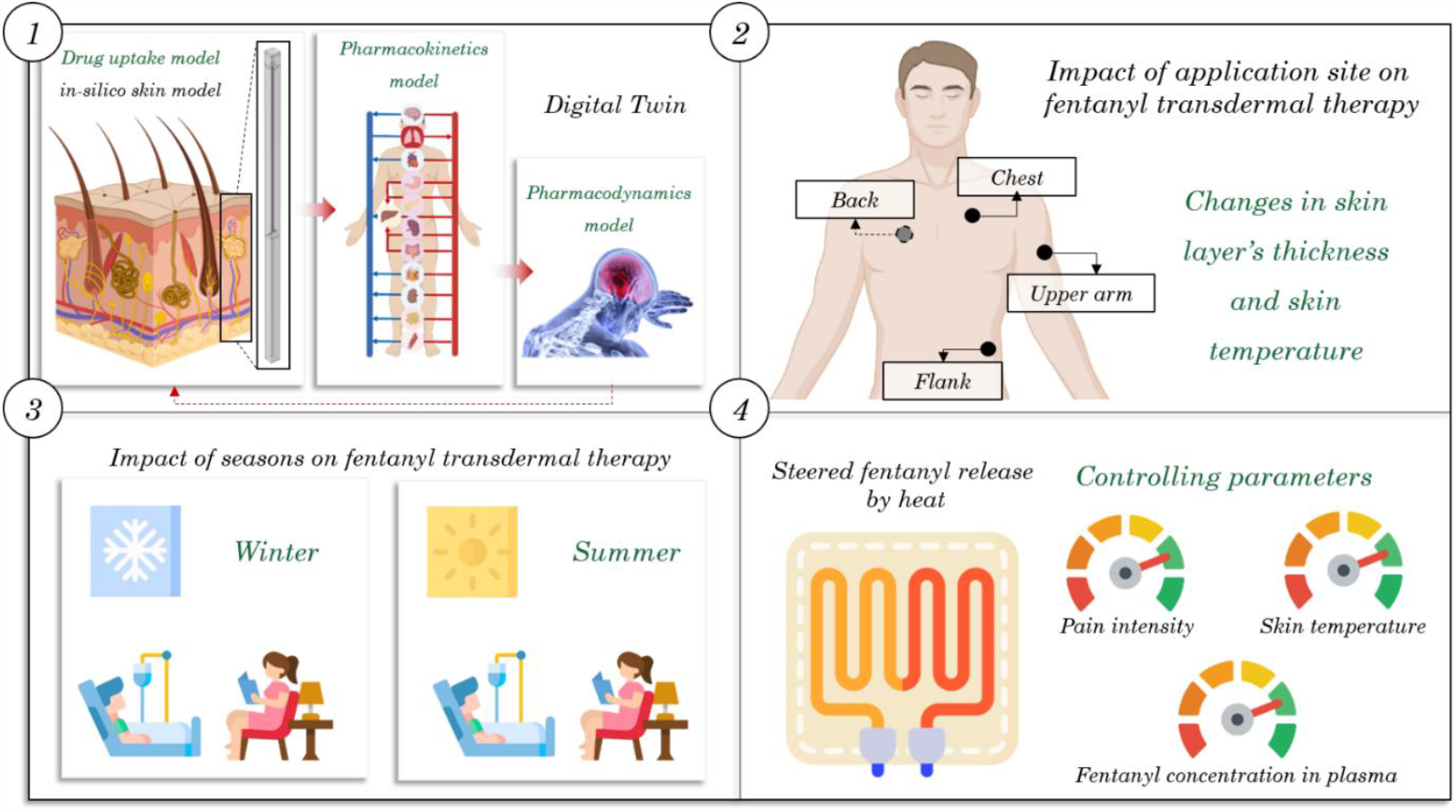
The overall structure of this study, which takes into account a detailed structure of the skin, the impact of body sites on drug uptake, the impact of ambient temperature on drug uptake, and finally, heat-enhanced delivery to control pain management therapy. (Created with elements from BioRender.com and www.flaticon.com)

### 2.1 Digital twin

#### 2.1.1 Drug uptake model for the skin

##### 2.1.1.1 Geometry

The geometry involved in the drug uptake model includes a patch, stratum corneum, viable epidermis, and dermal papillae, which provide a curved interface between the epidermis and dermis, dermis, capillary vessels, and hypodermis, which is shown in Figure 2. it should be noted for simplification the arterial capillary is connected directly to the venous capillary; however, the drug uptake is only considered via the venous end. The surface of this geometry is as wide as 70 μm, which is the same size as the surface area of each papilla. The drug uptake was calculated for the periodic model, and at the end, it was scaled up to the actual size of a fentanyl patch. The measurements regarding the geometry sectors are provided in Table 2.

**Figure 2.**
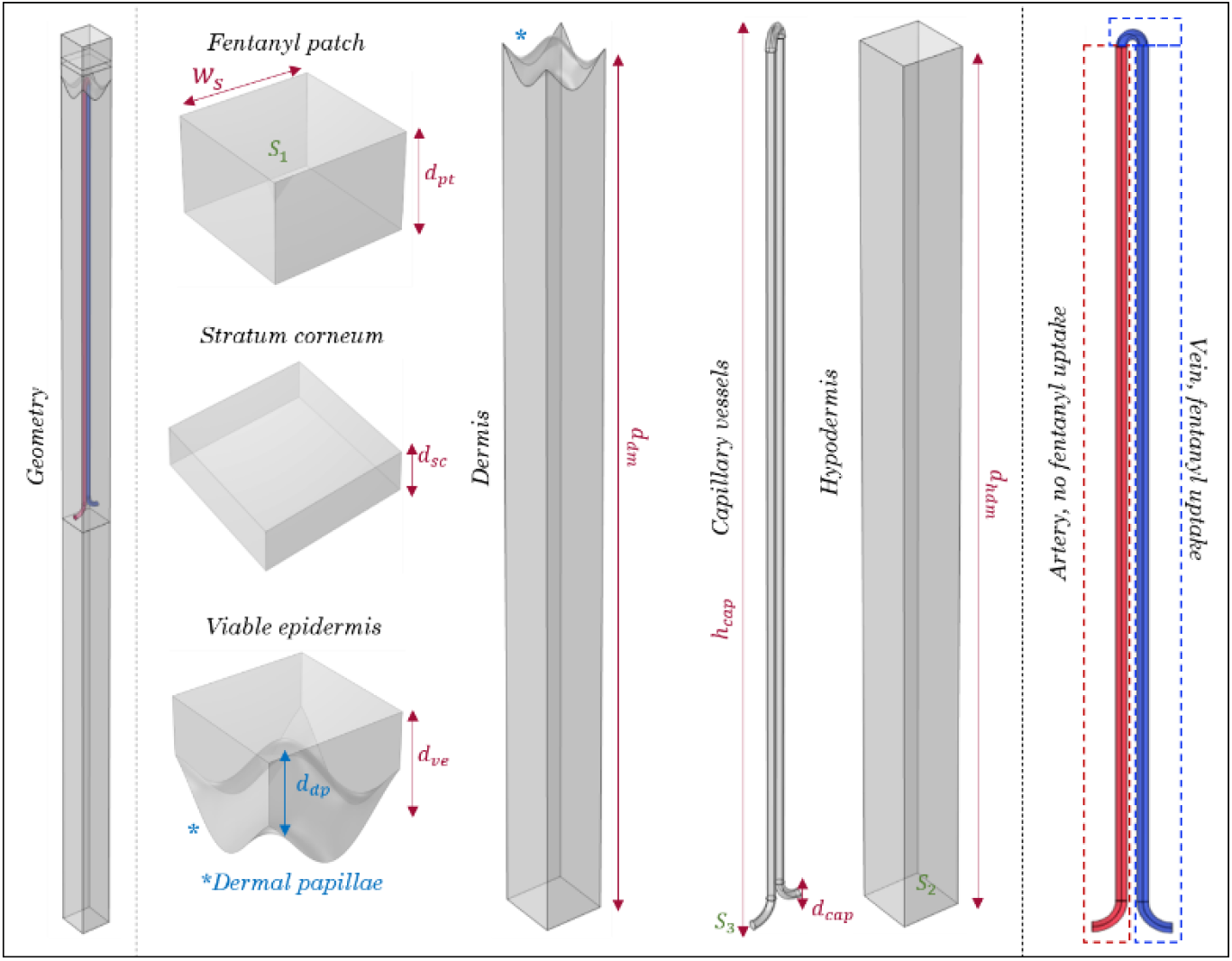
Geometry of fentanyl patch and skin layers included in the drug uptake model.

##### 2.1.1.2 Blood flow in the capillaries

Due to the small diameter of capillary vessels and low blood flow, the Reynolds number for flow in capillaries is 0.001, corresponding to creeping flow, an extreme case of laminar flow (G. E. Miller 2012). On the other hand, blood is a non-Newtonian flow, although it shows Newtonian behavior in many cases (Rahman and Haque 2012). However, the presence of red blood cells in the capillaries would reduce the accuracy of blood flow results from the Navier-Stokes model (Fullstone et al. 2015). Nevertheless, for simplification in this study, we assumed that blood in capillaries has a Newtonian behavior, and we implemented Navier-Stokes equation to model it (Siebert and Fodor 2009).

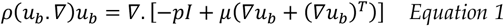

Here *ρ* [*Kg/m*^3^], *u*_*b*_ [*m/s*], *pI* [*Pa*], and *μ* [*Pa. s*] are blood density, blood velocity, pressure field, and dynamics viscosity, respectively. We assumed, for simplicity, that no plasma leaves and enters the capillary system; therefore, continuity is applicable to blood flow under our assumptions.

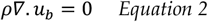

The following boundary conditions were applied for blood flow in the capillaries. The blood flow entered from surface S3 in Figure 2 with an average velocity of 0.065 cm/s based on the average blood flow velocity in the capillaries present in the dermis layer (Fagrell, Fronek, and Intaglietta 1977). We assumed that the blood flow in contact with the capillary wall (S4) has a 0 m/s velocity. The resting blood flow rate in the skin depends on several factors, such as temperature, age, humidity, and pressure (Petrofsky 2012; Vuksanović, Sheppard, and Stefanovska 2008). However, for simplicity, we assumed that the blood flow does not change over time and has thus steady states throughout the simulation.

##### 2.1.1.3 Diffusion process

Fick’s second law was implemented to monitor the fentanyl penetration from the patch through the skin layers and its uptake by the capillary network (Equation 3). The details of this model are provided in our previous studies (T. Defraeye et al. 2020; Thijs Defraeye, Bahrami, and Rossi 2021).

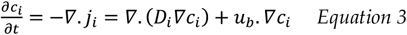

Where *c*_*i*_ *[kg/m*^*3*^*], j*_*i*_ *[kg/m*^*2*^.*s]*, and *D*_*i*_ *[m*^*2*^*/s]* are fentanyl concentration, fentanyl flux, and diffusion coefficient of fentanyl in domain *i*, respectively. These domains include patch, SC, viable epidermis, demis, capillaries, and hypodermis. However, solving the above equation is computationally demanding due to the different partition coefficients between different layers. Therefore, drug potential instead of drug concentration was used as it is continuous throughout the geometry. Drug potential is connected to drug concentration based on the partition coefficients.

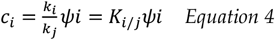

Where *k*_*i*_ *[-]* and *ψ*_*i*_ *[kg/m*^*3*^] are drug capacity and drug potential in domain *i*, respectively. *K*_*i/j*_ *[-]* is the partition coefficient at the interface of domain *i* and *j*.

#### 2.1.2 Pharmacokinetics model

As fentanyl is taken up by the bloodstream, it gets distributed throughout the body, metabolized in the liver, and excreted through the renal system. To track fentanyl’s journey in the body, especially its concentration in the plasma, we implemented a lumped Physiological-Based Pharmacokinetic (PBPK) modeling approach (Björkman 2003). Our model contains five compartments, each representing different organs based on their function. The first compartment is the central compartment, including blood circulation and the lungs (Equation 5). Fentanyl flux from the skin layers enters this compartment (Equation 6). The second compartment is the rapid-equilibrated compartment, including the heart, brain, skin, and kidneys (Equation 7) (Björkman 2003). The third compartment is the slow-equilibrated compartment, which consists of muscle, carcass, and adipose tissue (Equation 8) (Björkman 2003). The fourth compartment represents the gastrointestinal tract, involving the spleen, gut, and pancreas (Equation 5). Lastly, the fifth compartment is the hepatic compartment, where fentanyl metabolism occurs (Equation 6).

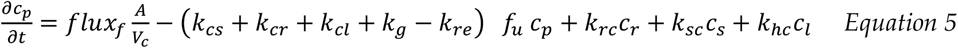

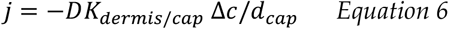

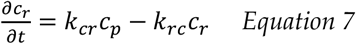

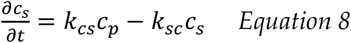

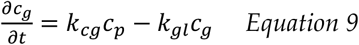

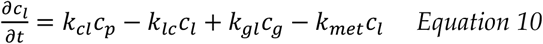

Where *c*_*i*_ [*Kg/m*^3^], *K*_*ij*_ [*s*^−1^], *f*_*u*_ [*%*], *flux*_*f*_ [*Kg/m*^2^*s*], *K*_*re*_ [*s*^−1^], and *K*_*met*_ [*s*^−1^] are fentanyl concentration in compartment *i*, first-order equilibrium rate constant from compartment *i* to *j*, unbound fraction of fentanyl, fentanyl flux to the blood circulation, renal clearance constant rate, and metabolism constant rate, respectively. *D, K*_*dermis/cap*_, Δ*c*, and *d*_*cap*_ are diffusion coefficient of fentanyl in dermis, partition coefficient of fentanyl between dermis and blood, concentration difference between dermis and capillaries, and thickness of capillary’s wall, respectively. The general overview of the pharmacokinetics model is shown in Figure 3.

**Figure 3.**
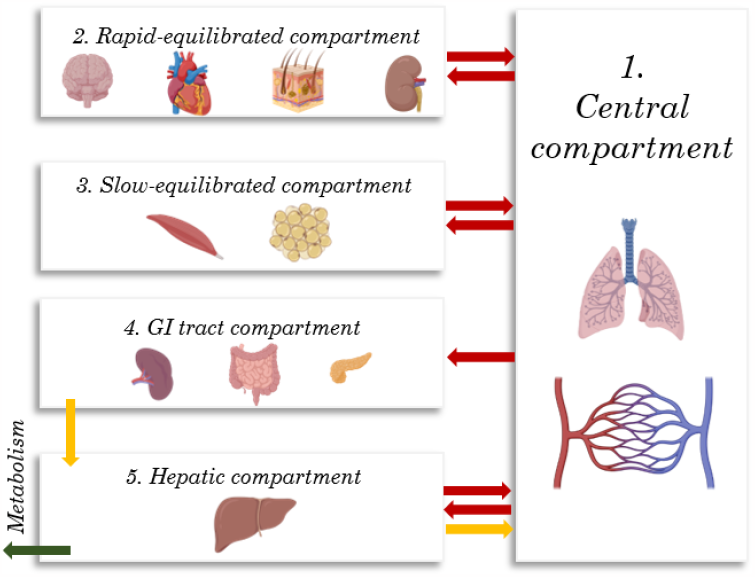
The compartments involved in the pharmacokinetics model. (Created with BioRender.com)

##### 2.1.2.1 Validation of drug uptake model with capillaries using plasma fentanyl concentration

In our previous studies, we validated fentanyl uptake from the patch through the stratum corneum and viable epidermis (T. Defraeye et al. 2020; Thijs Defraeye, Bahrami, and Rossi 2021) and fentanyl concentration was calculated by the pharmacokinetics model based on fentanyl uptake by skin model (Bahrami, Rossi, and Defraeye 2022). In this study, the skin model includes dermal papillae, dermis layer, capillary network, and hypodermis layer, which are not present in the prior validated drug uptake model. In order to validate the drug uptake model, we compared the evaluated fentanyl concentration in plasma by validated pharmacokinetics model to our previous model, in which the geometry included four blocks as a patch, SC, viable epidermis, and dermis. The thickness of the dermis was only a fraction of the whole dermis and was calculated based on the reported time lag of the fentanyl patch reported in the literature (Bahrami, Rossi, and Defraeye 2022). Additionally, we compared the simulated data with experimental data from the Marier et al. study (Marier et al. 2006). In their study, they had 24 male subjects aged 18 to 45 years old, with a BMI of 18 to 27 kg/m^2^ and a minimum weight of 60 kg. In this study, they implemented a fentanyl patch with a nominal flux of 50 μg/h on the upper arm of the volunteers. In order to evaluate the accuracy of the prediction of our model, we calculated the normalized root-mean-square deviation (NRMSD) and area under the curves (AUC).

#### 2.1.3 Pharmacodynamics model

There is a time lag between the concentration of fentanyl in plasma and the corresponding effect on pain relief. To take into account this time lag, an imaginary compartment, called the effect compartment, is considered. The concentration of fentanyl in the effect compartment by the biophase model is described in Equation 11 to connect the concentration in plasma to the therapeutic effect. (Felmlee, Morris, and Mager 2012).

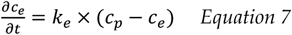

Where *c*_*e*_ [*Kg/m*^3^], *K*_*e*_ [*s*^−1^], and *c*_*p*_ [*Kg/m*^3^] are fentanyl concentration in the effect compartment, first-order equilibrium rate constant to the effect compartment, and fentanyl concentration in plasma, respectively. To calculate the pain intensity during fentanyl transdermal therapy, the sigmoid *E*_*max*_ model is implemented (Equation 8) (Felmlee, Morris, and Mager 2012). In *E*_*max*_ model, the therapeutic effect depends on the maximum response, the concentration related to half of the maximum response, the concentration of the drug in effect or central compartment, and the hill coefficient. The hill coefficient describes the steepness of the relationship between drug concentration and therapeutic effect (Felmlee, Morris, and Mager 2012).

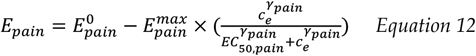

Where *E*_*pain*_, 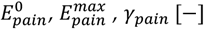, *γ*_*pain*_ [−], and *EC*_50,*pain*_ [*Kg/m*^3^] are pharmacological pain intensity (based on the VAS scale), the initial pain intensity, maximum possible pain relief, Hill coefficient, and concentration of half-maximum effect of fentanyl for pain relief.

#### 2.1.4 Impact of the application site on fentanyl uptake

The skin properties, such as the thickness of skin layers, vary all around the body. Therefore, applying the same fentanyl patch on different body sites will lead to a different outcome. Based on the SmPC (Summary of Product Characteristic) of the Duragesic® fentanyl patch, the fentanyl patch can be applied on the chest, upper arm, back, and flank. The thickness of skin layers in mentioned body sites is detailed in Table 2.

#### 2.1.5 Impact of temperature on fentanyl uptake

The diffusion coefficient of fentanyl in the skin is temperature-dependent. With rising temperatures, the diffusion coefficient of fentanyl exhibits a concurrent increase (Gupta et al. 1992). Consequently, a fentanyl dosage appropriate for a patient under one thermal condition could potentially result in over-or under-dosage for the patient experiencing a different thermal scenario. To predict this change, we used the well-known Arrhenius equation (La Count et al. 2020).

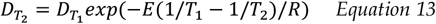

Where 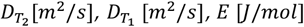, and *R* [*J/mol. K*] are diffusion coefficient at the temperature *T*_1_, diffusion coefficient at the temperature *T*_2_, the activation energy of diffusion, and the gas constant, respectively. Which here *T*_1_ is considered 33 °C, and 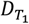 for the patch, SC, the viable epidermis (Thijs Defraeye, Bahrami, and Rossi 2021), and dermis (Karin Homber, Janay Kong, Sarah Lee 2008) were chosen based on our previous studies and the literature. Additionally, we assumed the diffusion coefficient of fentanyl in hypodermis is similar to the dermis.

##### 2.1.5.1 Heat transport modeling to calculate temperature distribution throughout the skin

As temperature impacts fentanyl uptake, we aimed to employ the characteristics of fentanyl thermally to enhance the fentanyl uptake by the skin in order to keep the fentanyl concentration in plasma and, eventually, the pain intensity in a desired range. To this end, we change the temperature at the surface of the fentanyl patch. We implemented the Pennes bioheat transfer equation to monitor the skin’s temperature distribution (Equation 14) (Nie, Zhang, and Song 2018).

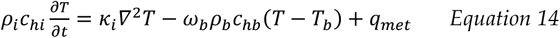

Where *ρ*_*i*_ [*Kg/m*^3^], *c*_*hi*_ [*J/Kg. K*], *κ*_*i*_ [*W/m. K*], *ω*_*b*_ [*m*^3^*/s. m*^3^], *ρ*_*b*_ [*Kg/m*^3^], and *c*_*hb*_[*J/ Kg. K*] are the density of skin layer *i*, the heat capacity of skin layer *i*, the thermal conductivity of skin layer *i*, blood perfusion rate, blood density, and the heat capacity of the blood, respectively. *T* [*K*], *T*_*b*_ [*K*], and *q*_*met*_ [*W/m*^3^] are temperature, blood temperature, and metabolic heat generation, respectively. Additionally, we considered the temperature at Surface S_2_ and S_3_ (in Figure 2) to be constant and equal to the average skin temperature.

##### 2.1.5.2 Skin temperature variability through different seasons

The ambient temperature changes considerably during different seasons, which, based on the patient’s activity states and clothing, may cause skin temperature and blood flow variations. In this study, we assume no changes happen in the thickness and diameter of the capillaries, and the changes in blood perfusion rate only appear in blood velocity. The changes in skin temperature can lead to changes in the fentanyl diffusion coefficient in skin layers and, eventually, the uptake of fentanyl. Therefore, the same fentanyl patch for one patient in different seasons may lead to different outcomes. In order to study this impact, we considered four scenarios, as listed in Table 1.

**Table 1.**
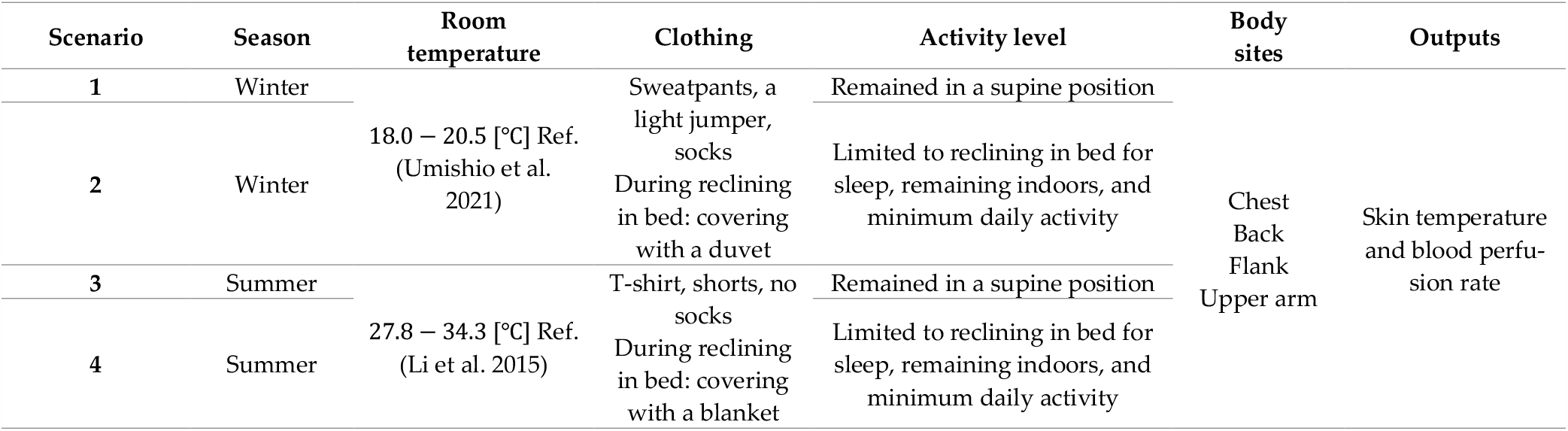
Summary of four scenarios considered for the impact of seasonal change in skin temperature and blood perfusion rate.

**Table 2.**
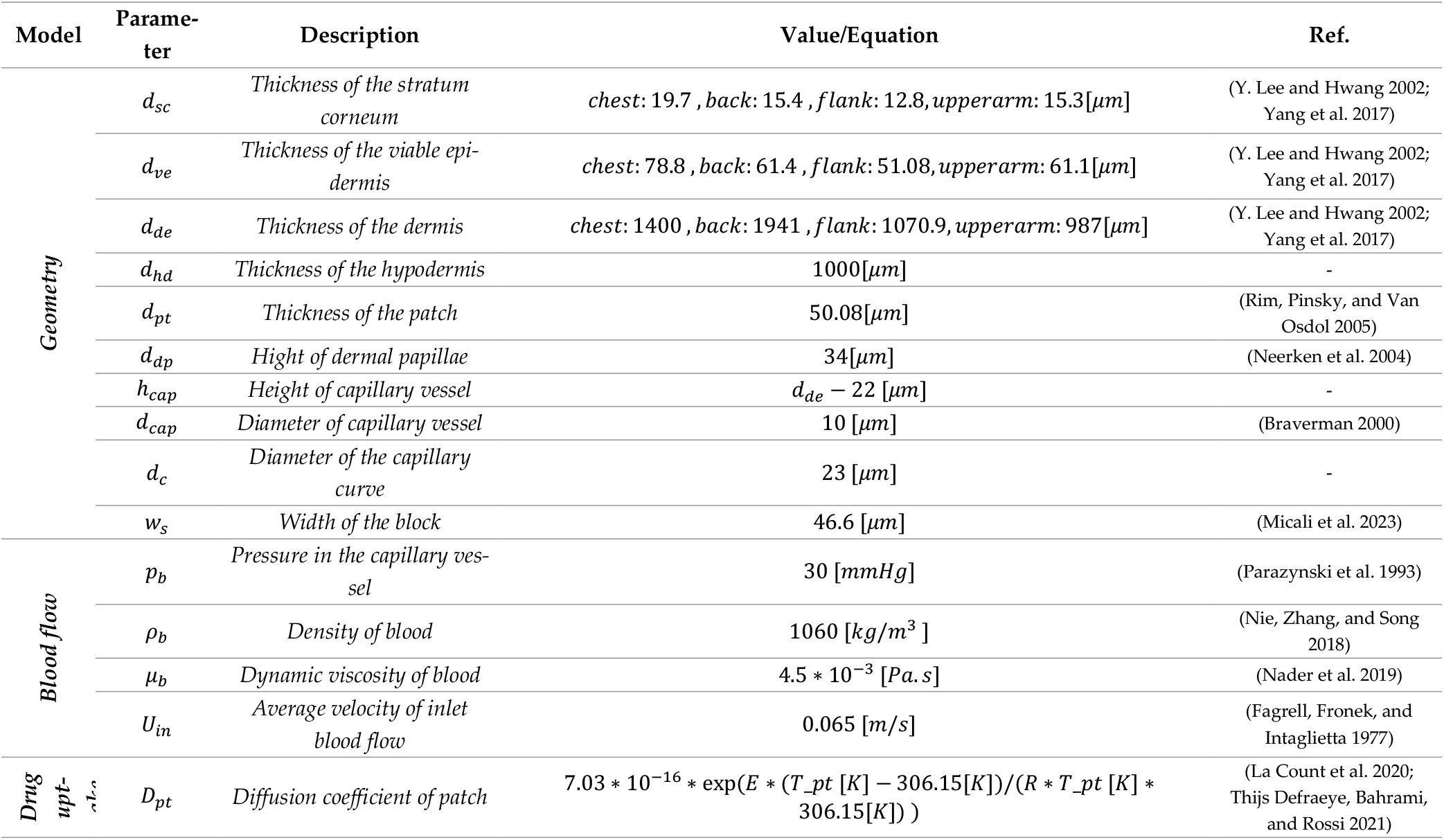
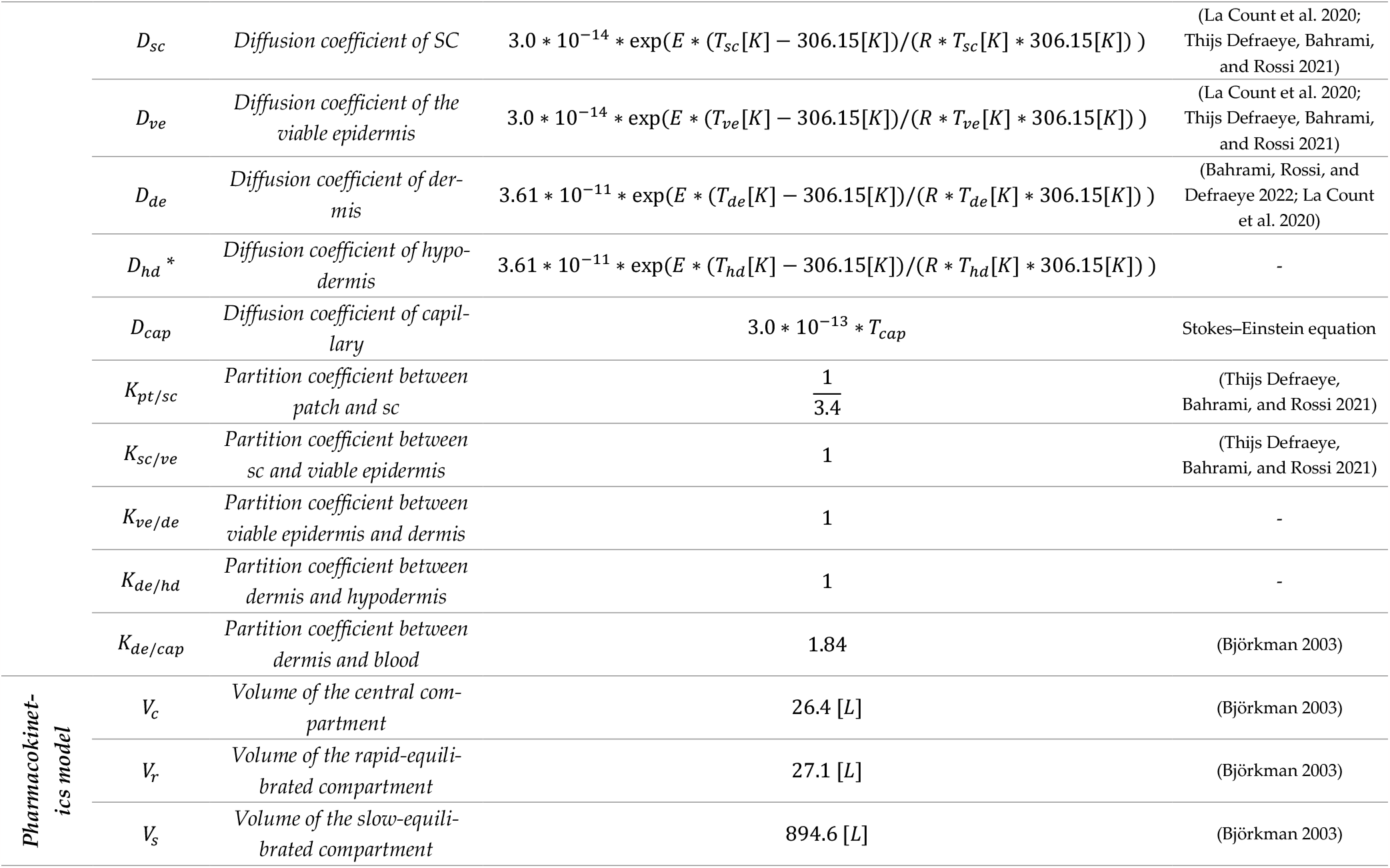
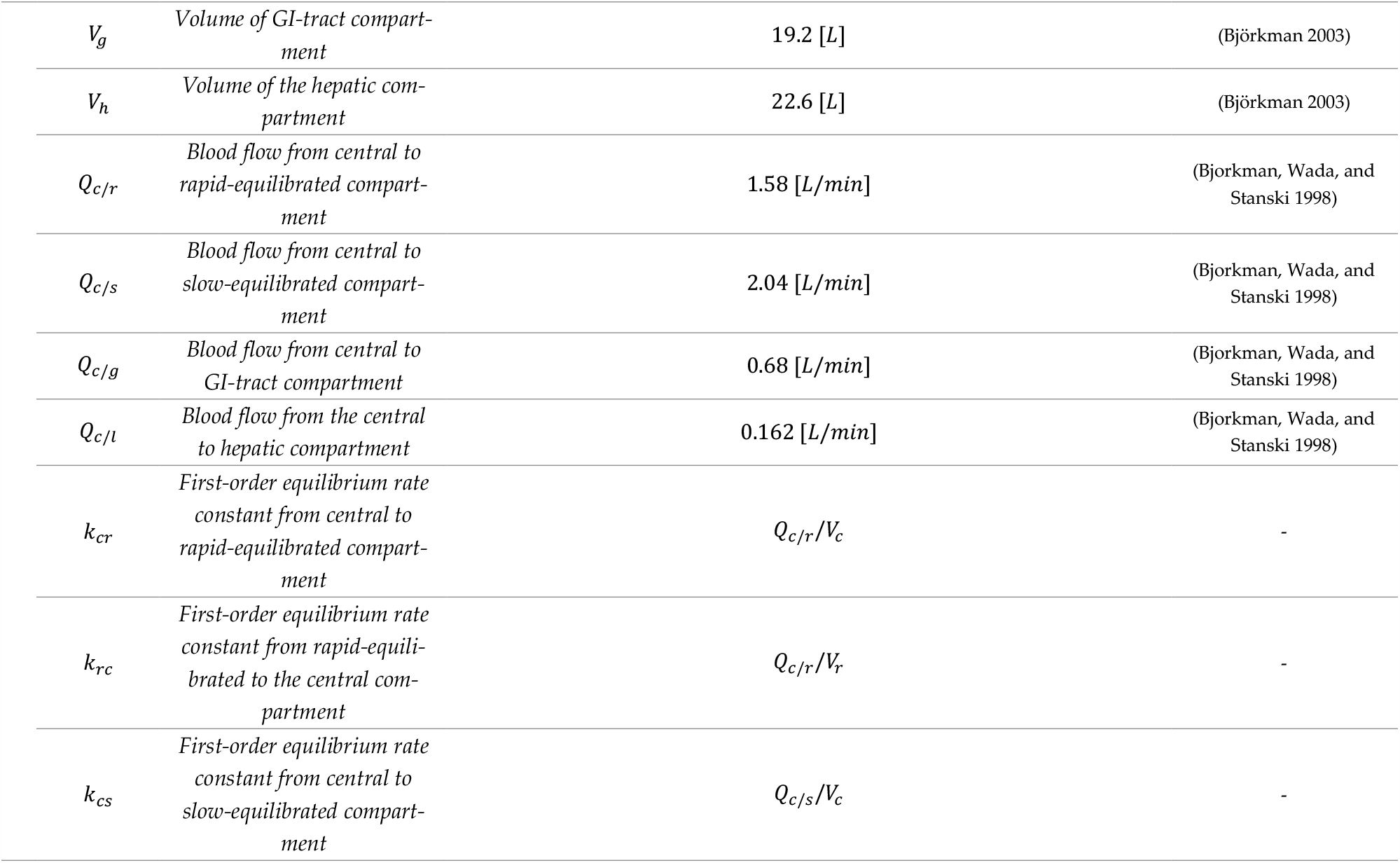
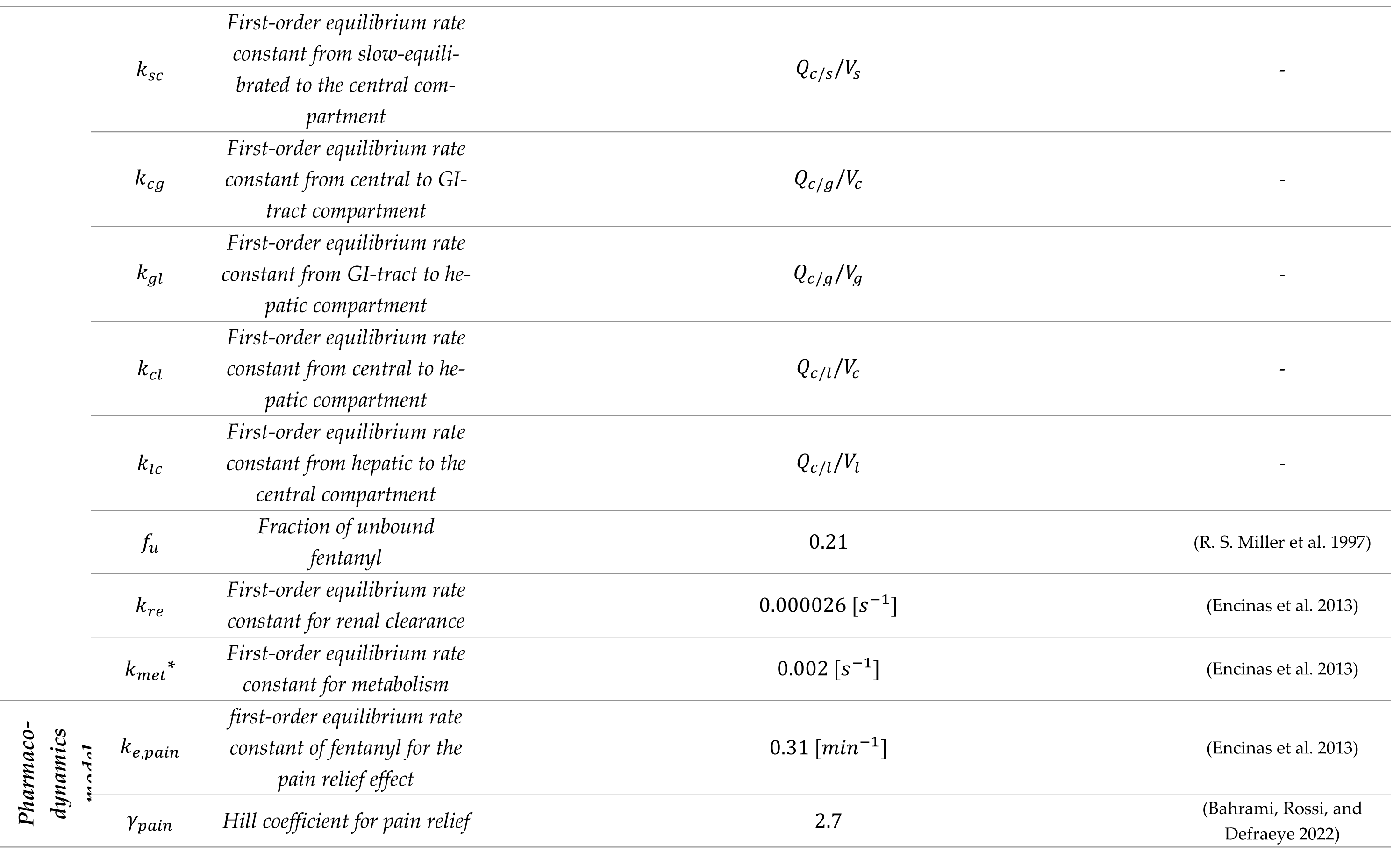
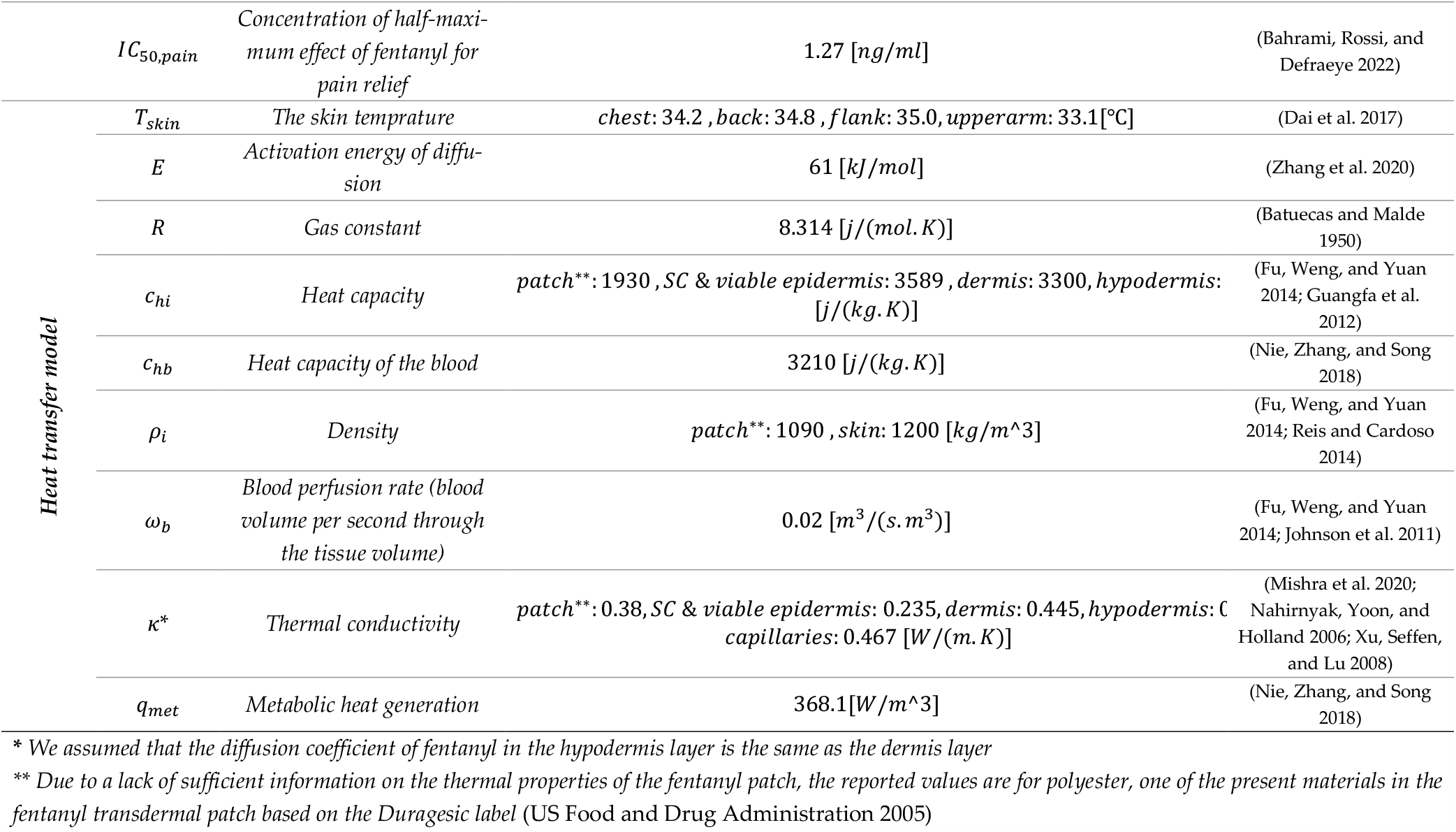
Parameters implemented in the developed digital twin.

The human thermoregulation model predicting human thermal response in an awake state to the environmental conditions at a given activity level and clothing situation (Fiala, Lomas, and Stohrer 1999, 2001) was used to simulate corresponding skin temperatures and blood perfusion rates at body location of interest. The model is based on the bioheat equation describing the energy balance of the human body and regression equations modulating skin blood perfusion rates, sweating, and shivering thermogenesis. The validity of the model was proven in several validation studies, including mean skin and core temperatures as well as local skin temperatures (Koelblen et al. 2018; Martínez et al. 2016; Psikuta et al. 2012). Clothing was addressed using data from a thermal manikin study (Fojtlín et al. 2019) to account for possibly the most realistic conditions of the thermal exposure.

The variation in skin temperature at different anatomical sites during the scenarios outlined in Table 1 is illustrated in Figure 4. As shown in Figure 4a&b, the skin temperature on the chest varies in summer from 33.9 °C to 35.2 °C for the inactive state, and for the active state, it varies between 33.8-35.4 °C. However, in winter, there is an almost 3 °C difference between the average skin temperature of the flank, chest, and back. Furthermore, as shown in Figure 4c&d, the skin temperature on the chest during winter for the inactive state varies from 31.9 °C to 33.8 °C and for the active state, from 32.4 °C to 34.0 °C. As skin temperature varies, so does the blood perfusion rate, as depicted in Figure 4e. Based on this result, the blood flow rate changes from 304% (vasodilation) of the thermoneutral state to 13.2% (vasoconstriction) when transitioning from the active state in summer to the inactive state in winter.

**Figure 4.**
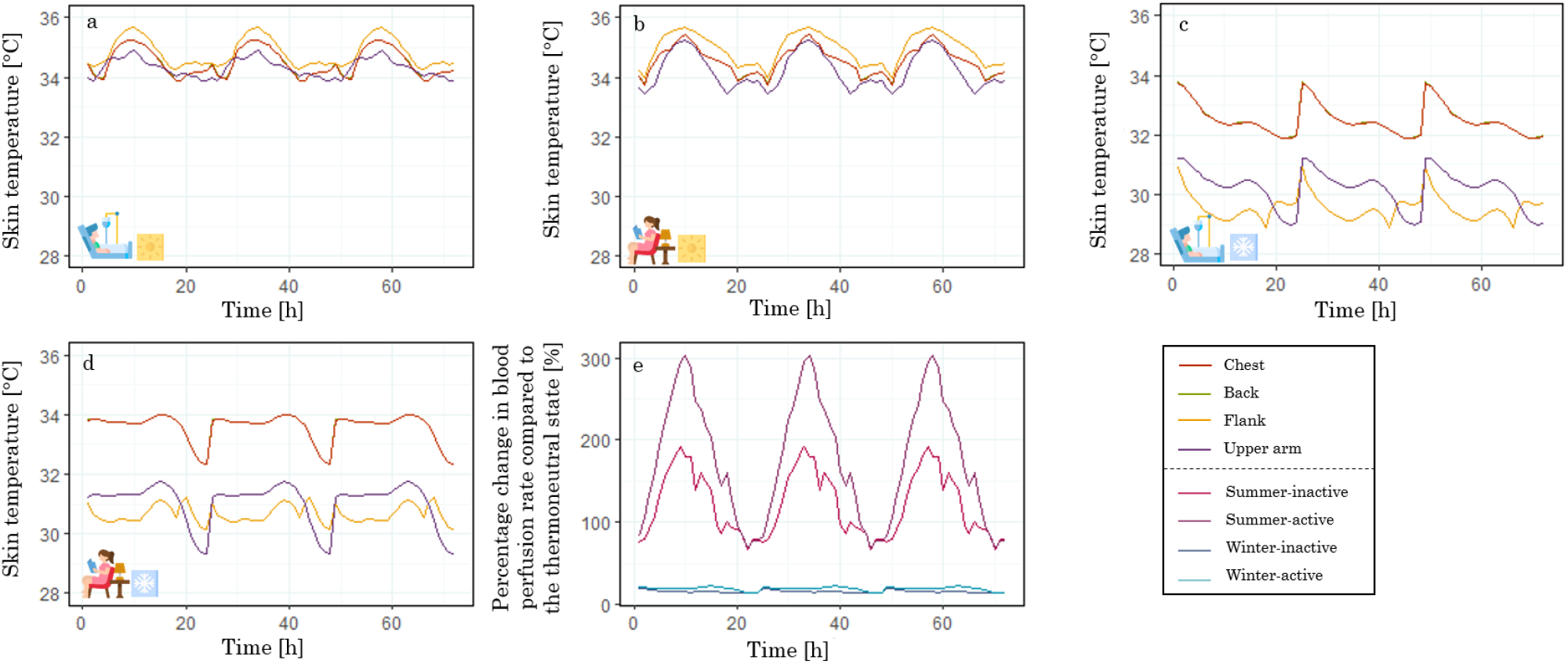
Skin temperature of the chest, back, flank, and upper arm during a: inactive state in summer, b: active state in summer, c: inactive state in winter, and d: active state in winter; e: the percentage blood flow rate compared to the base state during active and inactive states in summer and winter. (Created with elements from www.flaticon.com)

##### 2.1.5.3 Thermally-controlled fentanyl transdermal delivery

The diffusion coefficient of fentanyl in skin layers depends on the temperature. As the temperature increases, the diffusion coefficient of fentanyl; subsequently, the fentanyl flux from the skin and, eventually, the fentanyl concentration in plasma increases. In this study, we aimed to use this behavior of fentanyl in response to heat to steer the fentanyl transdermal therapy in order to keep the fentanyl concentration in plasma and its corresponding effects in a favorable range. To reach this end, we used the event interface of COMSOL Multiphysics to control the state of applying heat to the fentanyl patch to increase the temperature of the outer surface of the fentanyl patch to a maximum of 43.5 °C. Our goal was to maintain the fentanyl concentration below 2 ng/ml, pain intensity (VAS) below 3, and the stratum corneum temperature below 43 °C. However, in all cases, the fentanyl concentration remained below the 2 ng/ml limit, making this criterion unnecessary. The threshold of 43 °C for the temperature of SC was chosen since 43 °C is the highest temperature the skin can tolerate for a long time without disturbing the blood flow (Wienert, Sick, and Zur Mühlen 1983). Additionally, a higher temperature, like 44 °C, is the threshold of skin temperature for pain sensation (Greene and Hardy 1958), and tissue injury will occur if the skin stays at this temperature for at least six hours (Ong and Milne 2016). When the applying heat is off, we assumed the temperature of the outer surface of the patch switches back to 33 °C, which is a normal skin surface temperature (C. M. Lee et al. 2019). Therefore, the conditions below were applied.

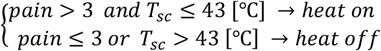

##### 2.1.5.4 Validation of temperature impact on fentanyl concentration in plasma

To assess the digital twin’s predictive accuracy in understanding heat’s impact on fentanyl uptake, we compared the calculated values by digital twin with the measured values in the study done by Shomaker et al. (Shomaker, Zhang, and Ashburn 2000). Their study involved six healthy volunteers aged 18 to 50, weighing 55 to 100 kg. Fentanyl patches with a nominal flux of 25 μm/h were applied on the upper right anterior chest, with experiments conducted both without heat and with 4 hours of heat application until the skin reached 41°C. In the Shomaker et al. study, the heat flux, the baseline skin temperature without heat, and changes in the temperature profile of the skin are not provided. Therefore, we assumed the skin temperature during no-heat duration to be the average skin temperature of the chest (34.2 °C (Dai et al. 2017)) and 41 °C while applying the heat. To evaluate the prediction performance of the digital twin in the impact of heat on drug uptake, NRMSD was calculated.

### 2.2 Estimation of model parameters

The value of implemented parameters used in blood flow, drug uptake, pharmacokinetics, pharmacodynamics, and temperature distribution model used in this study is provided in Table 2. The parameters implemented in the drug uptake model, pharmacokinetics model, and pharmacodynamics were validated in our previous studies (Bahrami, Rossi, and Defraeye 2022; Thijs Defraeye, Bahrami, and Rossi 2021).

### 2.3 Spatial and temporal discretization

Tetrahedral elements were employed to grid the skin layers and capillary networks, with the number and distribution of these grids selected based on the involved physics. Notably, the density of these grids increased at the interfaces. Due to variations in thickness at different body sites, the number of grids differed accordingly. For evaluating the blood velocity in the capillary network in the fluid flow model, the number of grids ranged from 47,503 to 55,837. In contrast, the number of grids for calculating the fentanyl flux through the skin layers varied between 1,811,291 and 3,184,822. This variation in the number of grids for each model was due to the changes in skin layer thickness on different anatomical sites. The simulations were conducted for a maximum of 72 hours (3 days), matching the application time of the fentanyl transdermal patches. The COMSOL model determines the time steps implicitly based on the BDF (Backward differentiation formula) method; however, for achieving high temporal resolution in the recorded data, we chose a time step of 0.1 h.

### 2.4 Numerical implementation and simulation

COMSOL Multiphysics version 6.1 was utilized to simulate various aspects, including blood flow in the capillaries, fentanyl penetration through the skin, fentanyl distribution and elimination in the body, fentanyl effects, and temperature distribution in skin layers. The solver scheme employed in this study was MUMPS (MUltifrontal Massively Parallel sparse direct Solver). A laminar flow module and stationary study were used for blood flow in the capillary network. Fentanyl penetration through the skin and temperature distribution in skin layers were modeled through partial differential equations (PDEs) and time-dependent studies. The pharmacokinetics model was established using boundary ordinary differential equations (ODEs) and time-dependent studies. The concentration of fentanyl in the effect compartment was calculated using the ODE interface, and the effects of the drug were analyzed using boundary probes and time-dependent studies. The event interface and time-dependent study were employed to control the heating state.

## 3 Results and discussion

### 3.1 Fentanyl uptake by the skin

#### 3.1.1 Validation of drug uptake model with capillaries using plasma fentanyl concentration

Implementing the information from Section **Error! Reference source not found**. on the subjects and the condition outlined in the Marier et al. study (Marier et al. 2006), we calculated the fentanyl concentration in plasma using our study’s developed digital twin. Based on the result shown in Figure 5, our previous study’s NRMSD (normalized root-mean-square deviation) is 0.15, and the current model is 0.1. Therefore, the current model predicted closer values to experimental data. The time to reach the maximum concentration (*t*_*max*_) for the current model is 36 h, which is the same as the experimental data; however, for our previous model, it is 23 h (only based on the recorded times, not the actual one). The maximum fentanyl concentration in plasma for experimental data is 1.7 ng/ml; for the previous model, it is 1.2 ng/ml (3% lower), and for the current model, it is 1.1 ng/ml (11% lower). The area under the curve for experimental data is 64.7 ng h/ml; for the previous model is 69.4 ng h/ml (7.7% more), and for the current model is 63.7 ng h/ml (1.5% less). To conclude, this new model with blood flow not only predicts the fentanyl concentration in the plasma closer to the experimental data and with a similar area under the curve but also predicts the *t*_*max*_ more accurately. However, compared to our previous model, the predicted maximum concentration of fentanyl in the plasma is 8% less than in experimental data.

**Figure 5.**
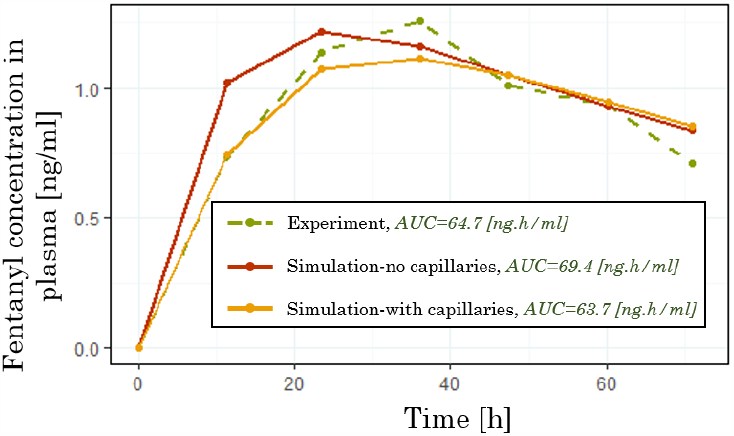
Fentanyl concentration in the plasma during 72 hours of applying one fentanyl patch with a nominal flux of 50 [μg/h]. (The green curve is based on results from the Marier et al. study (Marier et al. 2006), and the red curve is based on results from the Bahrami et al. study (Bahrami, Rossi, and Defraeye 2022)).

### 3.2 Impact of application site on fentanyl uptake

We virtually applied a fentanyl patch with a nominal flux of 50 *μm/h* on the chest, back, flank, and upper arm of a virtual patient with a body mass of 70 kg and age of 60 years old. These locations vary from each other based on their skin layers’ thickness and skin temperature, which values are provided in Table 2. Based on the results in Figure 6a, the main deviation in fentanyl flux from the path to different body sites occurs at the initial time. The main cause of this variation is the different skin temperatures, which lead to different diffusion coefficients. The fentanyl flux toward the capillary networks is shown in Figure 6b. Based on this result, the flux of fentanyl in different application sites is considerably different from each other; however, except for the patch on the flank, the fentanyl concentrations in plasma (Figure 6c) are very similar. The reason is that besides the flux, the capillary surface area that the fentanyl penetrates through differs in each of them, which is directly dependent on dermis thickness. For example, the fentanyl flux for the upper arm is higher compared to the back, which is 93% more, while the dermis thickness is 49% less. Therefore, the capillary surface area for fentanyl uptake in the upper arm is considerably less than in the back. Additionally, the maximum concentration of fentanyl in plasma (*c*_*max*_) and time to reach this maxmimum concentration (*t*_*max*_), the most two important pharmacokinetics parameters vary for different application sites. The *t*_*max*_when fentanyl patch is applied on the upper arm happens 10.3 h later than the flank, on the other hand, the *c*_*max*_ when the fentanyl patch is applied on the flank is 15.3% higher than the chest. Howevere, the difference in *c*_*max*_ when the patch is applied on the upper arm and the back is only 1.2% and 1.7% higher than the chest. As expected, the calculated pain intensity only when the patch is applied on the flank is considerably different from the other application sites. However, for none of the applications, the pain intensity reaches VAS 3, which is a medium pain intensity and the target outcome in this study. From these findings, applying the same fentanyl patch for the same patient on different anatomical sites can lead to different outcomes. Consequently, the effectiveness of a fentanyl patch in managing the pain on one anatomical site does not guarantee success on another site.

**Figure 6.**
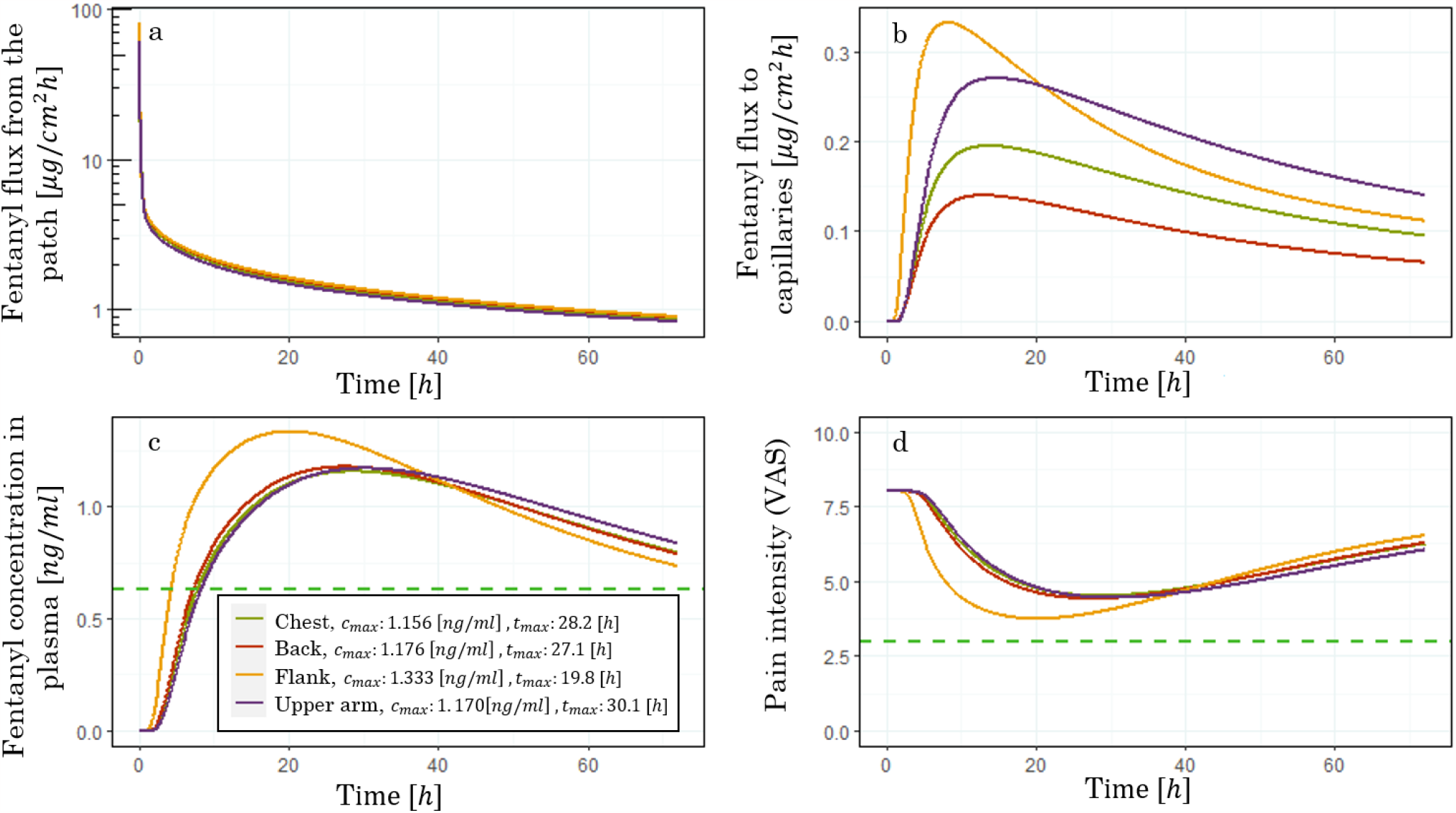
the impact of the application site, including chest, back, flank, and upper arm on a: fentanyl flux from the patch; b: drug uptake flux by capillaries; c: fentanyl concentration in plasma; and d: pain intensity

### 3.3 Thermal impact on fentanyl transdermal therapy

#### 3.3.1 Validation of temperature impact on fentanyl concentration in plasma

Based on the conditions and information on the subjects of the experiment in the Shomaker et al. study (Shomaker, Zhang, and Ashburn 2000) provided in Section 2.1.5.4, we calculated the fentanyl concentration in plasma with and without applying heat on the skin and the patch by using the developed digital twin. Figure 7a reveals that the digital twin accurately predicted fentanyl plasma concentration during the no-heat experiment, with an NRMSD of 0.12 and only an 8.4% lower area under the curve compared to the experiment. In the experiment with applied heat (Figure 7b), the NRMSD was 0.15, and the area under the curve for simulated data differed by only 1% from the experiment. However, the maximum heat-induced concentration was 22.7% lower than the experimental data for the simulated data. Focusing solely on the digital twin’s prediction of heat effects, the concentration difference between heated and nonheated experiments was examined for both experiment and simulation (Figure 7c). In conclusion, the model could predict the impact of heat on fentanyl concentration in the plasma with a favorable level of agreement, which led to an NRMSD of 0.15.

**Figure 7.**
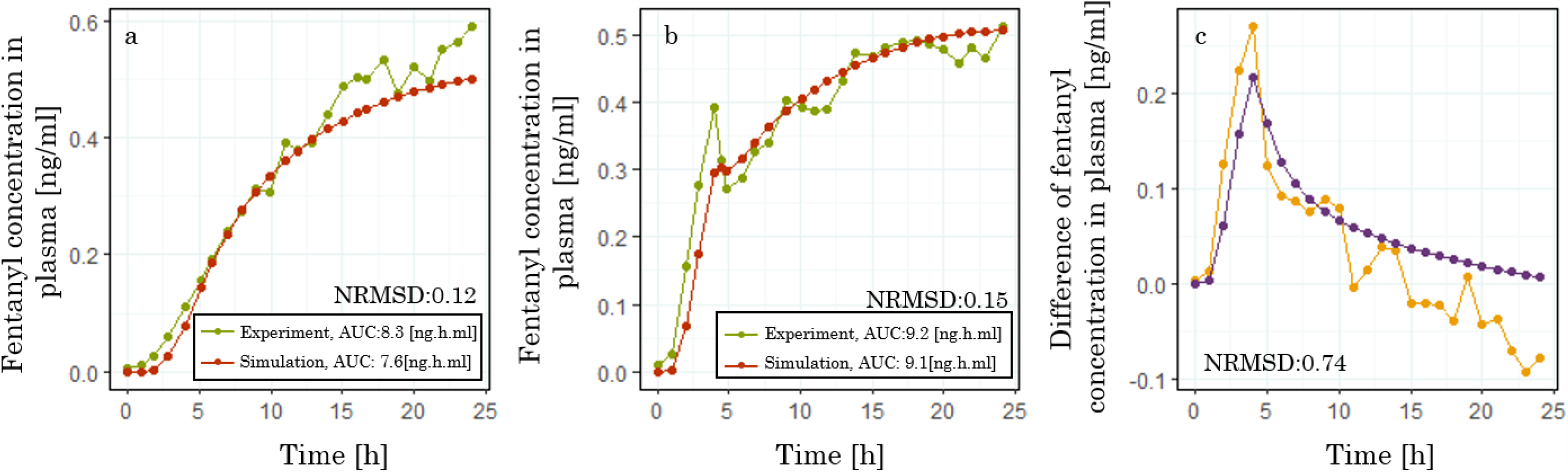
Validation of prediction of digital twin on the impact of heat on fentanyl concentration in plasma

### 3.4 Seasonal impact on fentanyl transdermal delivery

The ambient temperature and skin temperature fluctuate with the changing seasons, transitioning from winter to summer. Moreover, the patient’s condition, shifting from inactive to active, will impact skin temperature. Notably, changes in ambient temperature and activity state not only influence skin temperature but also affect the blood perfusion rate within the skin, as demonstrated in Figure 4, based on the conditions provided in Table 1. The results of implementing the calculated skin temperature and blood perfusion rate percentage into the digital twin for the fentanyl patch, with a nominal flux of 50 μg h^-1^ on the chest, back, flank, and upper arm, are shown in Figure 8. Based on this result, the maximum flux of fentanyl from the dermis layer to the capillaries increases by 1.8%, 2.6%, 11.8%, and 10.3% when transitioning from an inactive state in winter to active states in summer, with the patch applied on the chest, back, flank, and upper arm, respectively (Figure 8a,d,g,&j). Consequently, the maximum fentanyl concentration under the same conditions increases by 0.8%, 0.8, 1.5%, and 4.3% when the patch is applied on the chest, back, flank, and upper arm, respectively (Figure 8b,e,h,&k). As a result of variations in the concentration of fentanyl in the plasma, the minimum pain intensity under the same conditions increased by 0.9%, 1.1%, 2.1%, and 4.9% when the patch is applied on the chest, back, flank, and upper arm, respectively (Figure 8c,f,i,&l). Based on these results, the standard deviation for minimum pain intensity during these scenarios is five times more when the patch is applied on the upper arm than on the chest. This implies the higher impact of ambient temperature and activity when the patch is applied on the upper arm compared to the chest. Therefore, based on the condition described in Table 1, when the average room temperature drops from 31 °C to 19 °C, and the condition of the patient changes from active to inactive, the pain relief can be reduced by up to 4.9%. In cases of more extreme environmental variations, the impact on the outcomes of transdermal fentanyl therapy can be even more substantial. The total amount of released fentanyl in 72h for a fentanyl patch with a nominal flux of 50 μg/h during the mentioned thermal scenarios in Table 1 is shown in Figure 9. Based on these results, the total intake of fentanyl varies between different scenarios, such as the total amount of released fentanyl when the patch is applied on the upper arm during the active state in summer is 8.7% more than the inactive state in winter.

**Figure 8.**
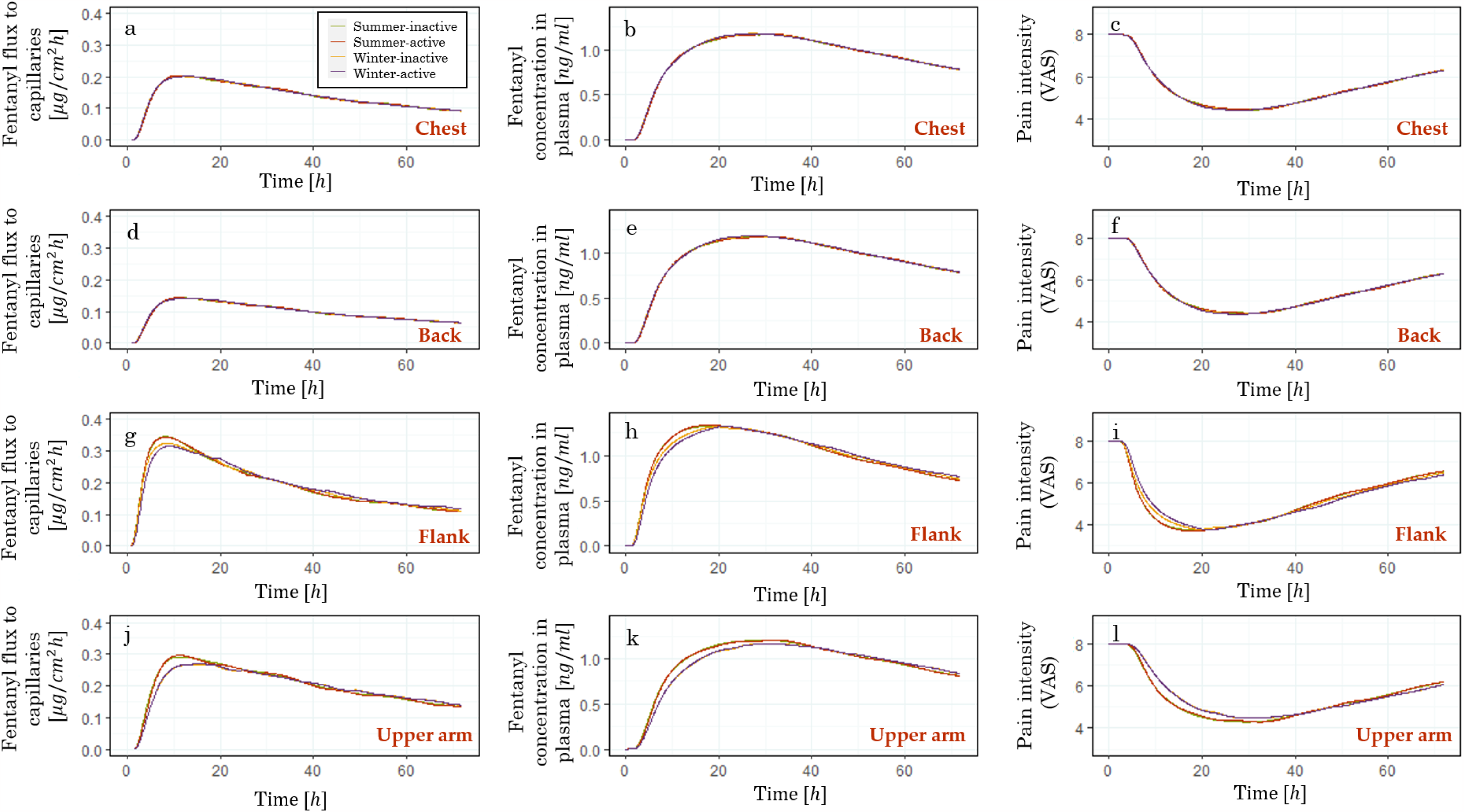
The impact of seasonal change and the activity of the patient on fentanyl flux to capillary (from a: chest, d: back, g: flank, and j: upper arm), fentanyl concentration in plasma (when the patch is applied on the b: chest, e: back, h: flank, k: upper arm), and pain intensity (when the patch is applied on the c: chest, f: back, i: flank, and l: upper arm)

**Figure 9.**
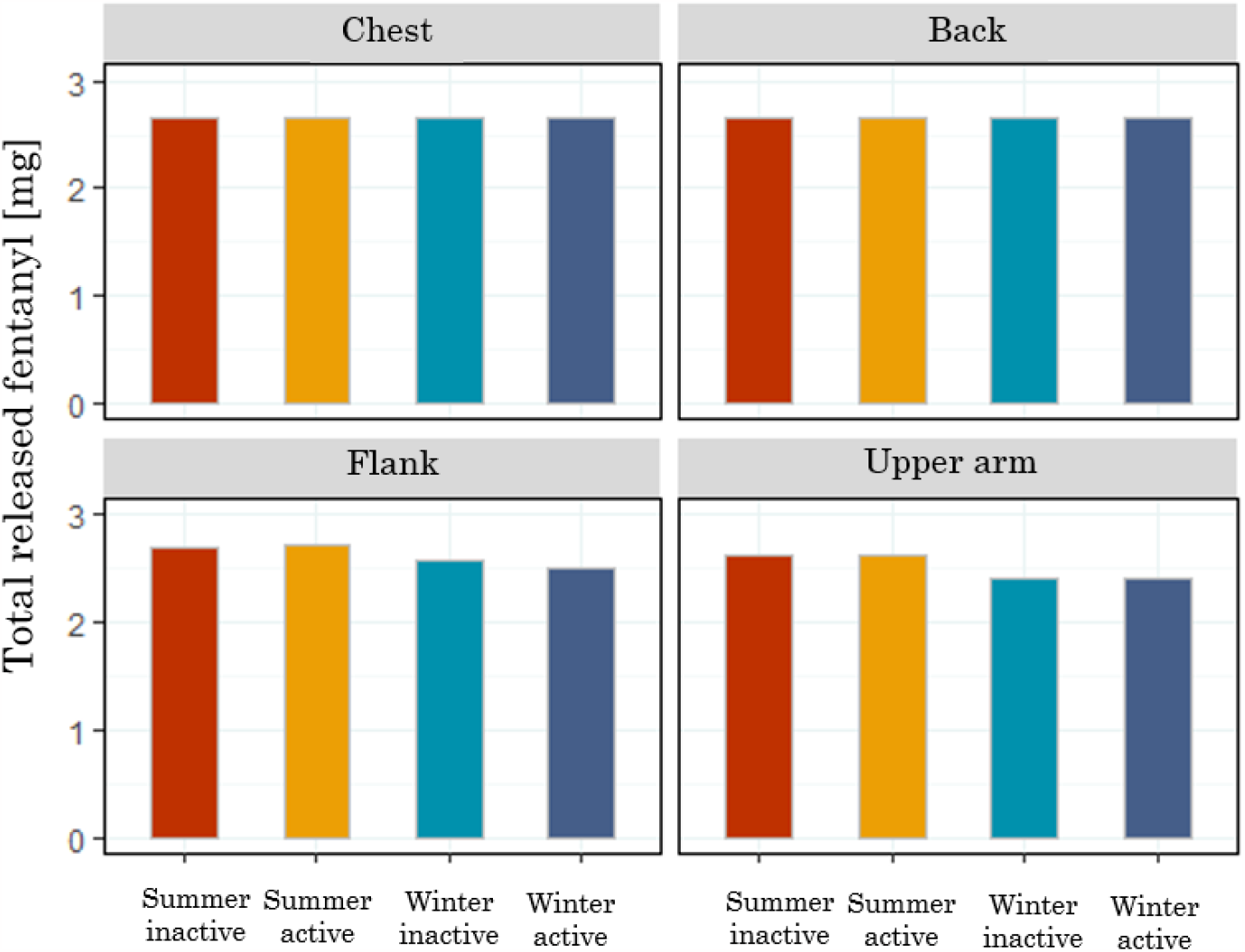
The total amount of released fentanyl from the patch with a nominal flux of 50 *μg/h* during different seasons and activity states.

### 3.5 Thermally enhanced fentanyl transdermal delivery

#### 3.5.1 Heat distribution in skin layers

The temperature distribution in skin layers and the patch is calculated based on the model provided in Section 2.1.5.1. In order to evaluate the impact of higher temperatures on the outer surface of the fentanyl patch on the diffusion coefficient of fentanyl in skin layers, we explored a range of temperatures from 33 °C, a normal skin temperature, to 43 °C, a safe, high temperature for the skin. Here we assumed the temperature at the bottome of hypodermis stay constant and equal to 33°C. As shown in Figure 10/a, by increasing the temperature at the outer surface of the fentanyl patch, the overall temperatures of the patch and skin layers increases; however, the changes decrease as the distance to the patch increases. As the patch surface temperature increases from 33 to 43 °C, the average temperature of SC changes from 33 to 42.5 °C, while the average temperature of hypodermis only changes from 33 to 33.3 °C. Following this, the diffusion coefficients of the patch, stratum corneum, viable epidermis, dermis, hypodermis, and capillary increase by factors of 2.1, 2.1, 2.0, 1.4, 1.0, and 1.0, respectively, when the outer surface of the patch is at a temperature of 43 °C, in comparison to when it is at 33 °C. Based on this result, by increasing the outer temperature of the fentanyl patch by 10 °C, the diffusion coefficient of fentanyl increases considerably, which may lead to higher penetration of fentanyl through the skin and eventually higher fentanyl concentration in plasma.

**Figure 10.**
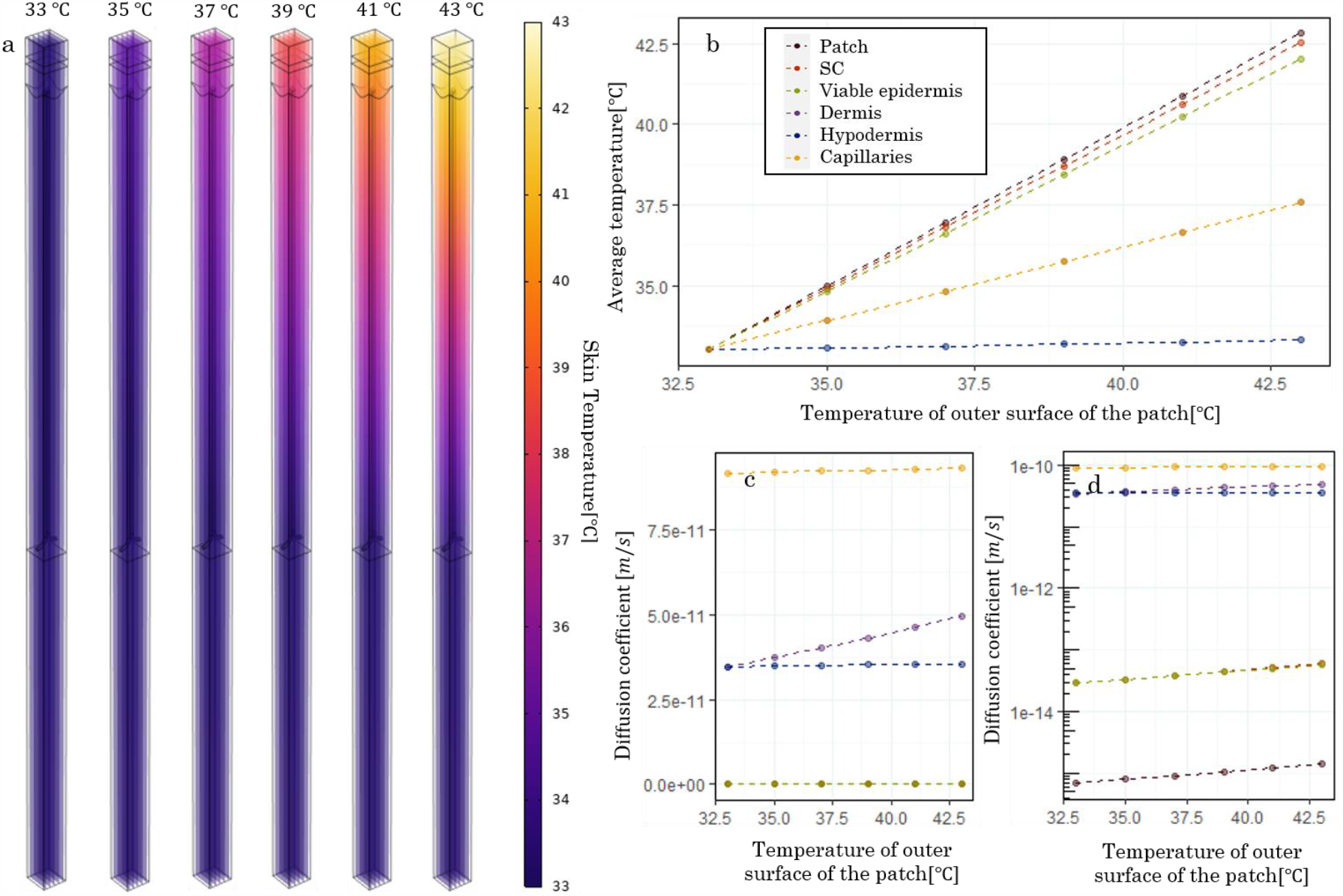
Temperature distribution in the skin and the patch during the application of different temperatures on the outer surface of the patch

##### 3.5.1.1 Thermally-controlled transdermal fentanyl delivery

Based on the results in Section 3.5.1, we aimed to enhance the fentanyl delivery by applying heat on the outer surface of the fentanyl patch. This could be, for example, done by designing a temperature-controlled wearable device or a heating garment and placing it on the patch. The heat application criteria are based on the conditions provided in Section 2.1.5.3. In Figure 11/a,b,&c, the fentanyl concentration in the plasma, pain intensity, and the average temperature of the stratum corneum are shown during 72 hours of fentanyl transdermal therapy with the nominal flux of 50[*μg/h*]. Based on this result, the virtual patient does not experience pain intensity below three throughout the therapy. However, after applying the heat on the surface of the fentanyl patch, the fentanyl concentration, as a result of the increase in the diffusion coefficient, increases (Figure 11/e). Due to the increase in the fentanyl concentration in plasma, the pain intensity for all the application sites drops below 3. Additionally, as shown in Figure 11f, the average temperature of the stratum corneum was kept under 43 [°C], and the heater’s temperature is shown in Figure 11g. The summary of thermally enhanced fentanyl delivery is brought in Table 3.

**Table 3.** Summary of the outcome of fentanyl treatment with and without applying heat on the outer surface of the fentanyl patch.

| Location | No heat |  |  | With heat |  |  |
| --- | --- | --- | --- | --- | --- | --- |
| | $c_{max}$ [ng/ml] | Average pain intensity (VAS) | Time without pain [h]* | $c_{max}$ [ng/ml] | Average pain intensity (VAS) | Time without pain [h]* |
| Chest | 1.16 | 5.5 | 0 | 1.60 | 4.4 | 14.6 |
| Back | 1.18 | 5.7 | 0 | 1.60 | 4.4 | 14.5 |
| Flank | 1.33 | 5.1 | 0 | 1.70 | 4.3 | 21 |
| Upper arm | 1.17 | 5.4 | 0 | 1.68 | 4.1 | 20.7 |
| SD | 0.08 | 0.18 | - | 0.05 | 0.12 | 3.6 |

\* The duration during which the pain intensity is below 3

**Figure 11.**
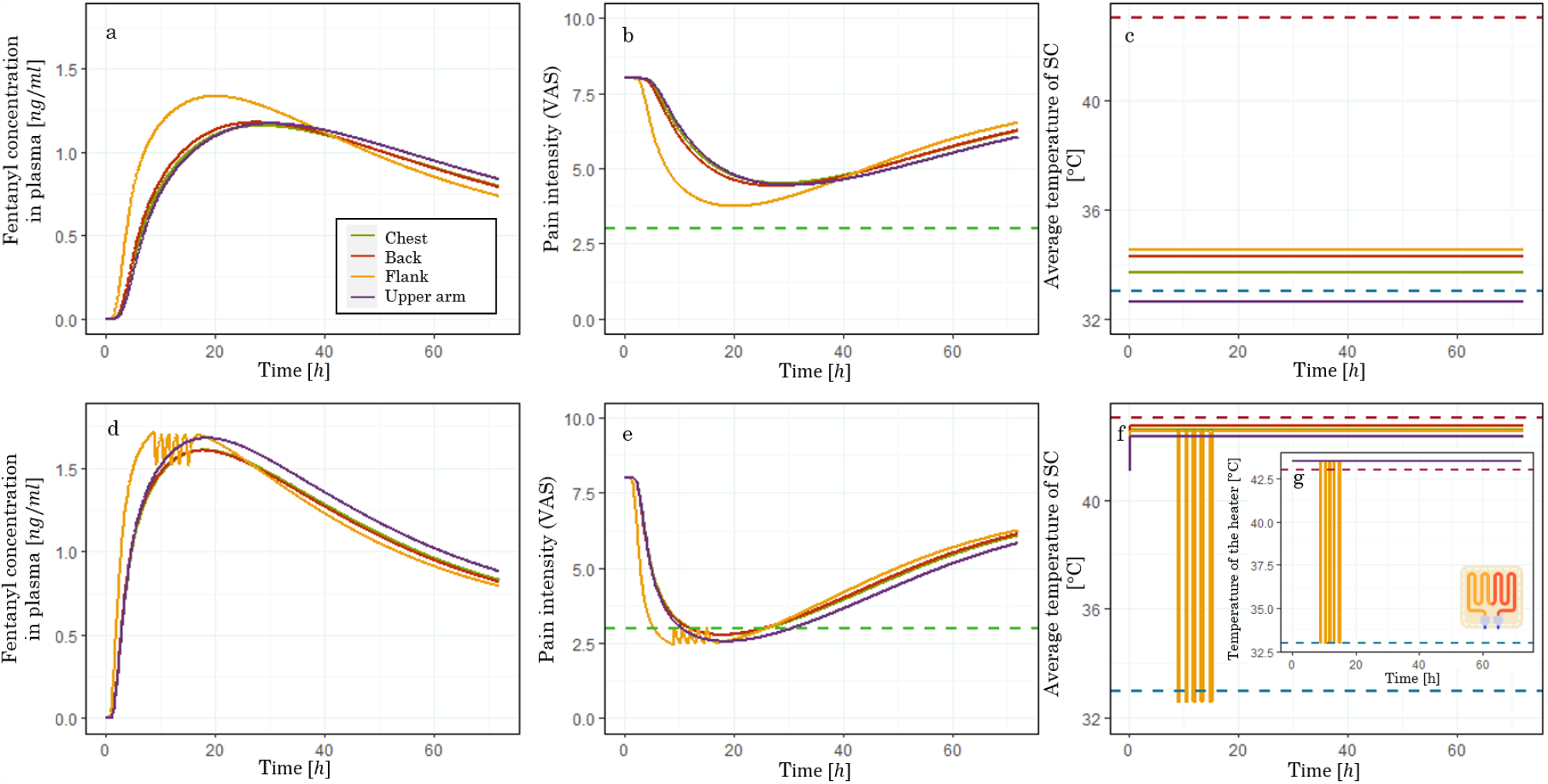
a: Fentanyl concentration in plasma; b: Pain intensity; c: Average temperature of stratum corneum during 72 hours of fentanyl transdermally therapy with applying no heat; d: Fentanyl concentration in plasma; e: Pain intensity; f: Average temperature of stratum corneum during 72 hours of fentanyl transdermally therapy with applying heat on the surface of fentanyl patch with a nominal flux of 50 μg/h. (The heathing icon is created with elements from www.flaticon.com)

Based on these results, which implemented controlled higher temperature, the digital twin is able to not only decrease the average pain intensity (between 0.8 to 1.3 units) and increase the time without pain but also reduce the deviation in fentanyl concentration and pain intensity between different application sites. Therefore, the digital twin was able to improve the outcome treatment while reducing the variability between different application sites by implementing higher temperatures on the fentanyl patch.

## 4 Outlook

Within the context of this research, we exclusively examined a virtual heater, which affects the temperature of the outer layer of the transdermal fentanyl patch. However, implementing an actual heater onto a transdermal patch would offer valuable insights into its flexibility in temperature adjustments and its limitations. This heater could be linked to a digital twin in order to control its activation and deactivation precisely, therefore providing a safe yet effective control release of fentanyl or other therapeutic substances.

In this study, the developed digital twin of the human body was implemented to monitor and steer transdermal fentanyl penetration through the skin, its distribution and concentration throughout the body, and finally, its therapeutic effect, pain relief. Nevertheless, fentanyl transdermal therapy is one of many examples of transdermal drug delivery systems that can be monitored and adjusted with the help of a physics-based digital twin. A similar approach by including the drug uptake model, pharmacokinetics model, and pharmacodynamics models can be implemented for other therapeutics drugs. Among suitable drugs for transdermal therapy, we can mention Estradiol for menopausal symptoms, Nicotine for smoking cessation, Lidocaine for post-herpetic neuralgia pain, Rotigotine for Parkinson’s disease, and Rivastigmine for dementia (Prausnitz and Langer 2008). Other types of transdermal patches with different drug uptake enhancers can also be investigated. Examples of these patches are iontophoresis, concavitational and cavitational ultrasound, electroporation, and microneedles.

In this study, a new skin model was introduced, which included capillaries with blood flow. This *in-silico* skin model included stratum corneum, viable epidermis, dermal papillae, dermis layer, hypodermis, and capillary network. We used this model to predict fentanyl penetration through the skin layers to reach the blood circulation system. Importantly, this virtual skin model can serve a broader purpose, extending to the assessment of the safety of cosmetics, toiletries, and topical products. This in-silico skin model, integrated with relevant kinetics throughout the body, offers essential insights that are beyond the capabilities of *in-vitro* studies without subjecting animals to the risks inherent in *in-vivo* tests. Therefore, this in-silico skin model holds promise as a valuable tool for physicians.

## 5 Conclusions

In this study, we developed a physics-based digital twin that includes drug uptake, pharmacokinetics, and pharmacodynamics models of fentanyl transdermal therapy. In this digital twin, an in-silico skin model incorporates stratum corneum, viable epidermis, dermal papillae, dermis, hypodermis, and capillary network. In order to validate the fentanyl uptake from this in-silico model, we compared the calculated fentanyl concentration with our previous model (Bahrami, Rossi, and Defraeye 2022) and experimental data from the Marier et al. study (Marier et al. 2006). The result showed that the novel skin model successfully predicted the fentanyl concentration in plasma (with NRMSD of 0.1), the area under the curve, and the time to reach the maximum concentration of fentanyl in plasma. However, it evaluated a lower maximum concentration of fentanyl in plasma compared to experimental data and the previous skin model.

Additionally, the impact of anatomical sites on fentanyl patch application was investigated. Due to the changes in skin layer thickness and skin temperature in different anatomical sites, the fentanyl uptake flux, fentanyl concentration, and pain intensity varied over different application locations. Based on our results, for a fentanyl patch with a nominal flux of 50 *μg/h*, the *c*_*max*_ for the patch applied on the chest, upper arm, and back is very similar and only varies for 1.7% and *t*_*max*_ varies for 3 h; however, the *c*_*max*_ for the flank is 15.3% more than the chest, and its *t*_*max*_ is 10.3 h less than the upper arm. This shows a high variation in the treatment outcomes between applying the fentanyl patch on the flank compared to the chest, back, and upper arm. Therefore, applying the same fentanyl patch on different body locations can lead to different therapy outcomes.

Furthermore, we conducted a study on the impact of seasonal changes and the patient’s activity state on skin temperature, blood perfusion rate, and, eventually, the fentanyl uptake rate. In this regard, we considered an active and inactive state for the patient, with clothing suitable for the seasons (winter and summer). Based on our findings, the transition from an inactive state in winter to an active state in summer resulted in an increase of up to 11.8% in fentanyl uptake flux by capillaries, an increase of up to 4.3% in fentanyl concentration, and an increase of up to 4.9% in the minimum pain level. With greater changes in the conditions throughout different seasons, the changes in the outcomes can be more drastic.

Ultimately, we took into account the impact of temperature on fentanyl diffusion in order to control the fentanyl release to reach the target outcome of fentanyl therapy, which we considered a pain intensity of 3 on the VAS scale. In this way, the average pain intensity (on the VAS scale) decreased from 5.44 to 4.12 from a no-heat condition to thermally enhanced delivery. The duration that the patient experienced mild pain intensity increased from 0h to 20.7 h. By implementing the thermally enhanced delivery of fentanyl, the standard deviation of c_max_ dropped from 0.08 to 0.05, and the average pain intensity standard deviation from 0.18 to 0.12. Therefore, not only did the thermally enhanced delivery of fentanyl improve pain relief, but it also reduced the variation in therapy outcomes throughout the treatment on different anatomical sites. Reducing therapy outcome variability leads to more predictable results, aiding physicians in patient treatment control.

## Data Availability

All data produced in the present study are available upon reasonable request to the authors.

## Acknowledgments

This study received financial support from the OPO Foundation (grant “Digital human avatars help tailor transdermal pain management (TREATME)”) and Margrit Weisheit Foundation and the Parrotia Foundation (grant titled “Digitale menschliche Avatare helfen bei der Anpassung der transdermalen Schmerztherapie in Echtzeit”). The funders had no role in the study’s design, data collection, analysis, interpretation, preparation of this article, or the decision to submit it for publication.

